# Integrative Genetic and Single-Cell Analysis Reveals Macrophages as Key Mediators Linking Aging and Osteoporosis

**DOI:** 10.64898/2026.09.09.26362574

**Authors:** Xin Li, Meng-Yuan Yang, Si-Rui Gai, Peng Wei, Zeng-Hui Gu, Ming-Yu Han, Yue-Zhou Wu, Jia-Sheng Yu, Wang-Jun Chen, Zhen-Rui Liao, Jia-Xuan Gu, Jia-Dong Zhong, Pian-Pian Zhao, Ke Zhu, Ching-Lung Cheung, David Karasik, Hou-Feng Zheng

**Author notes:** Corresponding: Dr Hou-Feng Zheng, M.D., PhD, Suzhou Laboratory of Precision Health and Data Science, the Second Affiliated Hospital of Soochow University, Suzhou, Jiangsu, China. These authors contributed equally (Xin Li, Meng-Yuan Yang).

## Abstract

Osteoporosis is a highly prevalent age-related disorder, and accumulating evidence suggests that it does not arise in isolation but is intrinsically linked to the aging process. Here, we employed integrative methods to systematically identify the relationship between aging and osteoporosis, and uncover the critical cell types and molecular regulators that mediate this association. Observational analysis of phenotypic data from UK Biobank and Mendelian Randomization analysis of summary-level statistics revealed that aging and osteoporosis are interrelated, acting as both causes and effects of each other. We identified macrophages as the key cell types mediating this association by integration of GWAS data with a comprehensively assembled single-cell transcriptomic atlas of bone remodeling. And the percentage of macrophages from bone marrow decreases with age. Additionally, shared genetic tools and weight co-expression network analysis were employed to uncover novel molecular regulators underlying this crosstalk. We identified *TGFB1* and a novel gene, *HNRNPUL1*, as key regulators that influence macrophages differentiation and function. In conclusion, our study provides observational and genetic evidence for a bidirectional relationship between aging and osteoporosis and unveils a macrophage-centric mechanism regulated by *TGFB1* and *HNRNPUL1*, offering new insights for age-related bone loss.

## Introduction

Osteoporosis is a prevalent skeletal disease characterized by decreased bone mineral density (BMD) and deteriorated bone microarchitecture, leading to an increased risk of fractures (Hendrickx et al. 2015; Rachner et al. 2011; Qian et al. 2021). As bone loss accelerates with age, aging is a major and causative factor for osteoporosis (Pignolo et al. 2021). Bone is a dynamic organ maintained by a regulated balance between bone formation and bone resorption, involving various cell types (Crockett et al. 2011; Bai et al. 2021). Osteoblasts, the bone-forming cells, produce organic bone matrix and aid in mineralization (Karsenty et al. 2009), while osteoclasts, the bone-degrading cells, dissolve bone mineral and enzymatically degrade extracellular matrix proteins (Teitelbaum 2007). With aging, this balance shifts negatively, leading to greater bone resorption compared to bone formation, ultimately resulting in decreased BMD and low bone mass (Demontiero et al. 2012; Josephson et al. 2019). Specifically, senescent bone marrow mesenchymal stem cells (MSCs) alter their proliferation and differentiation, favoring the adipogenic lineage, which impairs bone formation (Corrado et al. 2020; Cai, Xiong, et al. 2023). Additionally, factors secreted by senescent immune cells and other cell types accumulate in the bone marrow, further contributing to osteoporosis (Hou et al. 2024; Li, Xiao, et al. 2021). Conversely, emerging evidence suggests that osteoporosis, a local disorder, can accelerate global aging. For instance, the conditional knock-out of bone marrow osteocytes not only causes osteoporosis but also shortens the lifespan in mice (Ding et al. 2022). Young osteocytes secrete extracellular vesicles that can ameliorate cognitive impairment and the pathogenesis of Alzheimer’s disease in APP/PS1 mice (Jiang et al. 2022). Bone remodeling is energetically expensive and involved metabolism also affects whole-body metabolism (Lecka-Czernik et al. 2025). These findings suggest a close link, and potentially a bidirectional causality, between aging and osteoporosis. It’s therefore essential to simultaneously research osteoporosis and aging. Understanding the cell types and molecular mechanisms underlying the link between local bone disorder and systemic aging will bring new insights into both osteoporosis and aging.

Current research has primarily focused on either the process of aging or osteoporosis individually. Genome-wide association studies (GWAS) are an unbiased method to understand the genetic basis of traits by identifying single nucleotide polymorphisms (SNPs) linked to disease-relevant phenotypes (Zhu et al. 2021). Bone mineral density estimated by heel quantitative ultrasound (eBMD) and fracture risk are widely used to screen osteoporosis (Gonnelli et al. 2005; Kemp et al. 2017; Trajanoska et al. 2018). GWAS studies on eBMD and fracture risk have been performed to study osteoporosis, providing summary-level statistics that systematically represent osteoporosis (Trajanoska et al. 2018; 23andMe Research Team et al. 2019). Since aging clocks harness omics data with machine learning models to build measures of biological age, summary-level GWAS statistics on aging clocks like GrimAge also show the opportunity to systematically study aging (Rutledge et al. 2022; Levine et al. 2018). Given the representations of aging and osteoporosis, methods in computational biology, such as Mendelian Randomization (MR), LDSC and shared genetic tools that utilize GWAS summary data of various traits, are capable of systematically studying osteoporosis and aging at the same time (Zhao et al. 2024; Zhu et al. 2018). Expanding on GWAS, strategies that integrate GWAS with single-cell RNA-sequencing (scRNA-seq) data have been proposed to identify trait-associated cells or cell types, thereby enhancing the resolution and biological interpretability of GWAS findings (Jia et al. 2022; Jagadeesh et al. 2022; Yazar et al. 2022; Zhang et al. 2022). However, the application of such approaches to study bone aging is hampered by the lacking of a comprehensive single-cell atlas encompassing all cell types involved in bone remodeling. Bone remodeling-associated osteoblasts and osteoclasts are relatively rare compared to other cell types, such as immune cells or hematopoietic cells in bone marrow. Moreover, osteoclasts are large, multinucleated cells formed through the fusion of monocytes/macrophages (Crockett et al. 2011; Haacke et al. 2025), making it technically challenging to capture all cell types involved in bone aging with a simple sequencing library construction.

In this study, analyses of phenotypic data and genetic data were employed to systematically investigate the causality between osteoporosis and aging. We integrated different single-cell datasets containing various bone remodeling cell types to obtain a more comprehensive bone remodeling cell type atlas using deep learning-based methods. Then, we conducted a single-cell GWAS (scGWAS) to identify key cell types involved in bone aging. Shared genetic tools helped identify novel genes that may regulate bone aging.

## Materials and Methods

### Phenotypic Data

Individual-level phenotypic data were obtained from the UK Biobank (Application 41376), following the granting of data access (Xia et al. 2022; Zhu et al. 2022; Qian et al. 2025). From an initial cohort of 439,982 participants, we retained 278,164 samples with complete information on heel bone mineral density (Heel BMD), fracture status, and age. The specific UK Biobank data field codes used to define these traits are listed in Supplementary Table 1.

### GWAS summary statistics and cis-eQTL summary statistics

Three Genome-Wide Association Study (GWAS) summary-level datasets related to osteoporosis were downloaded from GEFOS Consortium (http://www.gefos.org/), which are GWAS for eBMD (426,824 European samples) (23andMe Research Team et al. 2019), fracture (53,184 European fracture cases and 373,611 European controls) (23andMe Research Team et al. 2019) and fracture (ALLFX) (37,857 fracture cases and 227,116 controls; predominantly European descents) (Trajanoska et al. 2018). All samples involved in association with eBMD and fracture are white British individuals, while GWAS for fracture (ALLFX) is a meta-analysis containing 25 cohorts from Europe, United States, east Asia and Australia. We obtained two summary-level datasets for aging, GrimAge (34,710 European participants) (McCartney et al. 2021) and mvAge (effective n = ∼1.9 million European participants) (Rosoff et al. 2023), available at https://datashare.ed.ac.uk/handle/10283/3645 and https://zenodo.org/record/7926323. GrimAge is one of the advanced methylation aging clocks, and the results of association analysis using GrimAge could be used to represent aging (Rutledge et al. 2022; Levine et al. 2018). mvAge used a multivariate approach, combining healthspan, lifespan, extreme longevity, frailty, and epigenetic aging to assess aging individually, which significantly extended the sample size (Rosoff et al. 2023). By reversing the frailty and epigenetic aging to generate positive correlations with other variables, mvAge effectively serves as an anti-aging or senescence resistance variable (Rosoff et al. 2023). Summary-level statistics from mvAge were subsequently used to represent aging, when employing approaches that do not incorporate the direction of effect, such as scGWAS. Detailed information on summary-level statistics is provided in Supplementary Table 2.

Full cis-eQTL summary statistics was download from eQTLGen consortium (https://eqtlgen.org/cis-eqtls.html) (Võsa et al. 2021).

### Single-cell transcriptomics datasets

We included five single-cell transcriptomics datasets in this study: (1) a bone marrow niche organization dataset (Baccin et al. 2020) and osteoclastogenesis dataset (Tsukasaki et al. 2020), used to integrate a landscape containing vital bone remodeling cell types; (2) a mouse bone marrow of different age dataset (The Tabula Muris Consortium et al. 2018) and heterochronic parabiosis dataset (Ma et al. 2022), employed to investigate age-related changes in macrophage abundance; and (3) a human bone marrow single-cell transcriptomics dataset (Bandyopadhyay et al. 2024), serving as external validation (Supplementary Table 3).

### Analysis of phenotypic data

Following the download of phenotypic data, we retained 278,164 samples with complete information on heel bone mineral density (BMD), fracture status, and age. Outliers exhibiting heel BMD values exceeding three standard deviations (SD) from the mean were excluded. Subsequently, the relationship between age and heel BMD was modeled using Generalized Additive Models (GAM) via the ggplot2 R package (version 3.5.2). Kaplan-Meier survival analysis was performed using the survival R package (version 3.6.4).

### Mendelian Randomization analysis

Mendelian Randomization (MR) analysis was employed to infer causal relationships between exposures and outcomes using genetic variants as instrumental variables (Qian et al. 2026; Guan et al. 2025; Zhao et al. 2019). Instrumental variables (IVs) were selected based on sample size and the amount of SNPs to ensure strong exposure associations, with linkage disequilibrium (LD) parameters adjusted accordingly (Supplementary Table 4). All IVs demonstrated robustness with F-statistics > 10. After harmonizing the IVs with outcome data, we manually excluded SNPs strongly associated with the outcome using the ‘GWAS Catalog’ and ‘PhenoScanner’. Additionally, MR-PRESSO (Verbanck et al. 2018) was employed to filter out outlier SNPs.

To infer causality, we employed four primary methods: inverse variance-weighted (IVW) (Johnson 2012), Weighted median, MR-Egger (Bowden et al. 2015) and MR-PRESSO. A *P-value* of less than 0.05 indicated a significant causal association between the exposure and outcome. To ensure the robustness of the findings, several sensitivity analyses were conducted. First, Cochran’s Q test was used to evaluate consistency across different SNP effects (Bowden et al. 2017), with a *P-value* < 0.05 indicating significant heterogeneity. Second, horizontal pleiotropy was evaluated using both the MR-Egger regression intercept (where a deviation from zero indicates bias) and the MR-PRESSO framework. Third, a “leave-one-out” analysis was performed, where each SNP was sequentially excluded to re-estimate the causal effect, allowing for the assessment of whether any single SNP significantly influenced the overall effect estimate. All MR analyses were conducted using R (version 4.4.2), supplemented by the TwoSampleMR (version 0.6.8) (Hemani et al. 2018; Hemani et al. 2017) and MRPRESSO (version 1.0).

### Single-cell transcriptomics data analysis

For heterochronic parabiosis dataset, raw sequencing data were downloaded and processed using CellRanger (version = 7. 1. 0) to generate count tables. Count tables for other datasets were directly downloaded. The count tables and metadata were then loaded and combined using Python package Scanpy (version = 1. 11. 0) (Wolf et al. 2018). Cells with fewer than 200 detected genes and genes not expressed in at least 3 cells were removed. Cells have high levels of mitochondrial genes were also removed. Then the data was normalized using size factor normalization so that every cell has 10,000 counts, followed log transformed. Principle component analysis (PCA), neighborhood graph and clustering using Leiden method were subsequently performed.

Highly differentially expressed genes (DEGs) in each cluster were calculated as marker genes using the Wilcoxon rank-sum test implemented in the Scanpy package. For bone marrow niche organization dataset and osteoclastogenesis dataset, cell types were manually annotated by comparing marker genes to those provided in the original studies and Cell Taxonomy database (https://ngdc.cncb.ac.cn/celltaxonomy/). Automated cell type annotation was performed for the mouse bone marrow aging dataset and the heterochronic parabiosis dataset using the Omicverse package (Zeng et al. 2024) (version 1.6.8), while the human bone niche dataset was annotated using CellTypist (Domínguez Conde et al. 2022) (version 1.7.1) with default parameters.

To integrate the bone marrow niche organization dataset and the osteoclastogenesis dataset, the count tables were concatenated together using Scanpy. An initial integration was performed using scVI, a method based on hierarchical Bayesian model implemented with a deep neural network (Lopez et al. 2018). The model parameters were set as follows: n_latent=30, max_epochs=400, train_size=0.9, and batch_size=128. It’s then further refined using scANVI (Xu et al. 2021), a semi-supervised model that leverages cell type knowledge for a subset of the cells in the dataset to infer the states of the remaining cells. The entire integration was carried out using python package Scvi (version 1.2.0) (Gayoso et al. 2022). To further assess integration efficiency, we employed additional batch effect correction methods, including BBKNN, ComBat, and Harmony. Specifically, Harmony was executed using the harmonypy package (version 0.2.0), while BBKNN and ComBat were performed using functions implemented in the scanpy package.

### Combination of GWAS summary statistics with single-cell transcriptomics using scGWAS

To infer the cell types in which disease-associated genes manifest and to construct cellular modules that imply disease-specific activation of different processes, we combined GWAS summary statistics with single-cell RNA data using scGWAS (Jia et al. 2022). We first prepared the average expression profiles for each cell type. Then, we mapped SNPs from GWAS to genes using MAGMA (De Leeuw et al. 2015). The scGWAS analysis was performed with default settings (module_socre=penalty, r_include=0.1 and r_exclude=0.05).

### scWGCNA and cellular communication analysis

To mitigate the sparsity inherent in single-cell sequencing data, we aggregated similar cells into ‘metacells’ using the K-nearest neighbors (KNN) algorithm implemented in the scikit-learn package (version 1.5.2) (Pedregosa et al. 2011; Morabito et al. 2023). Subsequently, were identified and removed via hierarchical clustering with a cutting height threshold of 125. Weighted gene co-expression network was then constructed using pyWGCNA package (version 2.2.0) (Rezaie et al. 2023). We identified gene modules correlated with two key phenotypes: macrophage percentage and age. Finally, functional enrichment analysis for the significant gene set was conducted using pyWGCNA with default parameters; modules with a p-value < 0.05 were considered statistically significant. Cellular crosstalk between different cell types was calculated using CellphoneDB (Garcia-Alonso et al. 2022) python package (version 5.0.1) by leveraging CellphoneDB database of interacting molecules with single-cell transcriptomics data.

### Shared genetic analysis and colocalization analysis

We constructed conditional quantile-quantile (QQ) plots to visualize polygenic enrichment for eBMD/mvAge. ConjFDR was then used to identify SNPs jointly associated with eBMD and mvAge (Smeland et al. 2020). A *P-value* of < 0.05 was thought to be significant. Local genetic covariance analysis was performed using SUPERGNOVA (Zhang et al. 2021). The summary-level statistics were converted into standard format that LDSC understands using munge_sumstats.py included in LDSC package(Schizophrenia Working Group of the Psychiatric Genomics Consortium et al. 2015). The reference panel used was obtained from 1000 Genomes Project.

To infer multiple independent causal variants, we performed a colocalization analysis using the coloc.signals function from the coloc (version 5.2.3) R package with defaults prior probability (Wang et al. 2020). The method cond was selected, and no default p12 was specified in the software, so it’s assigned a value of 5×10^−4^. Signals were defined as colocalizing if the posterior probability of shared association signals (P4) was greater than 0.8.

## Results

### A bilateral relationship between aging and BMD/fracture

After selecting individual-level phenotypic data for 278,164 participants from the UK Biobank, we employed generalized additive models and Kaplan-Meier survival analyses to investigate the relationship between bone mineral density (BMD)/fracture risk and aging. Linear regression results indicated that heel BMD declines with age in both sexes; however, the rate of decline accelerates in females after age 50 (Figure 1A). Conversely, Cox regression analysis revealed that individuals with a history of fracture or lower heel BMD exhibited reduced survival rates (Figure 1B). These observational findings suggest a bidirectional relationship between aging and BMD/fracture risk.

**Figure 1.**
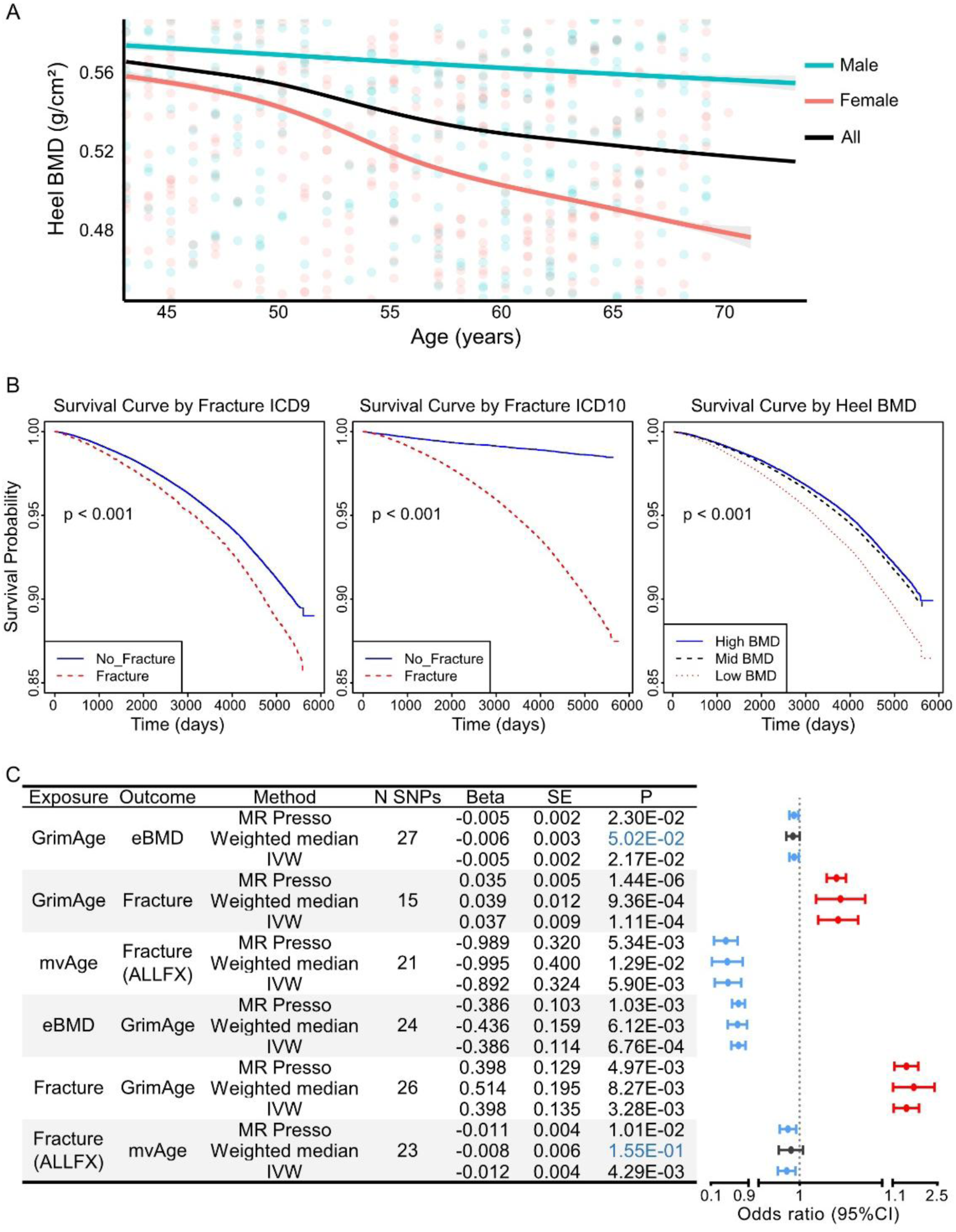
The potential association between aging and osteoporosis. (A) Regression models illustrating a gradual decline in heel bone mineral density (BMD) with age in both sexes after 45 years; (B) Cox regression results demonstrating lower survival rates in subjects with prior fractures or reduced BMD; (C) Causal effect estimates between aging and osteoporosis based on GWAS summary statistics. Significant causality (*P* < 0.05) was observed across MR-PRESSO, IVW, and Weighted median methods.

To further elucidate this link, Mendelian Randomization (MR) analyses was employed and the results from inverse variance weighted (IVW), weighted median, and MR-PRESSO methods consistently supported significant bidirectional causality, implying a molecular interplay between these traits. Specifically, using IVW estimates as the primary metric, higher genetically determined GrimAge (aging) was associated with lower eBMD (OR = 0.96, 95% CI = 0.95-0.97) and higher fracture risk (OR = 1.03, 95% CI = 1.01-1.05). The mvAge (anti-aging) was causally associated with a lower fracture risk (OR = 0.43, 95% CI = 0.22-0.82) (Figure 1C). Conversely, genetically determined fracture had a causal impact on GrimAge (OR = 1.49, 95% CI = 1.49-1.94) and mvAge (OR = 0.98, 95% CI = 0.97-0.99). Higher genetically determined eBMD was associated with lower GrimAge (OR = 0.68, 95% CI = 0.54-0.85) (Figure 1C).

To evaluate the robustness of the MR results, several sensitivity analyses were conducted. Results from leave-one-out analysis suggest that no SNP significantly altered the estimates (Supplementary Figure 1). Furthermore, after removing outlier instrumental variables (IVs) using MR-PRESSO, no significant heterogeneity was observed among the genetic instruments. Assessments for horizontal pleiotropy, including MR-Egger regression intercept tests and the MR-PRESSO global test, revealed no evidence of significant pleiotropy in any analysis (all *P* > 0.05, Supplementary Figure 1).

### Macrophage is a key cell type in BMD and aging susceptibility

Two single-cell transcriptomics datasets (Supplementary Figure 2) were employed to test the associations between cell type and trait, with one dataset containing cell type like immune cells, MSCs, osteoblasts differentiated from MSCs and others (Baccin et al. 2020), and the other primarily comprising cell types involved in osteoclast differentiation (Tsukasaki et al. 2020) (Supplementary Figure 2). Both datasets were loaded and integrated using a variational autoencoder (VAE)-based unsupervised learning method and then refined with further training on cell types (Neutrophils and B cells) shared by both datasets. Validation showed that shared populations (B cells and neutrophils) co-clustered effectively, while osteoclasts aligned with their progenitor monocytes from the other dataset (Figure 2A, Supplementary Table 5). Although traditional methods like ComBat and Harmony achieved higher aggregate scores, they failed to merge the datasets effectively, leaving identical cell types from different sources segregated. These results indicate that deep learning methods are essential for capturing the non-linear relationships within this data (Supplementary Figure 3).

**Figure 2.**
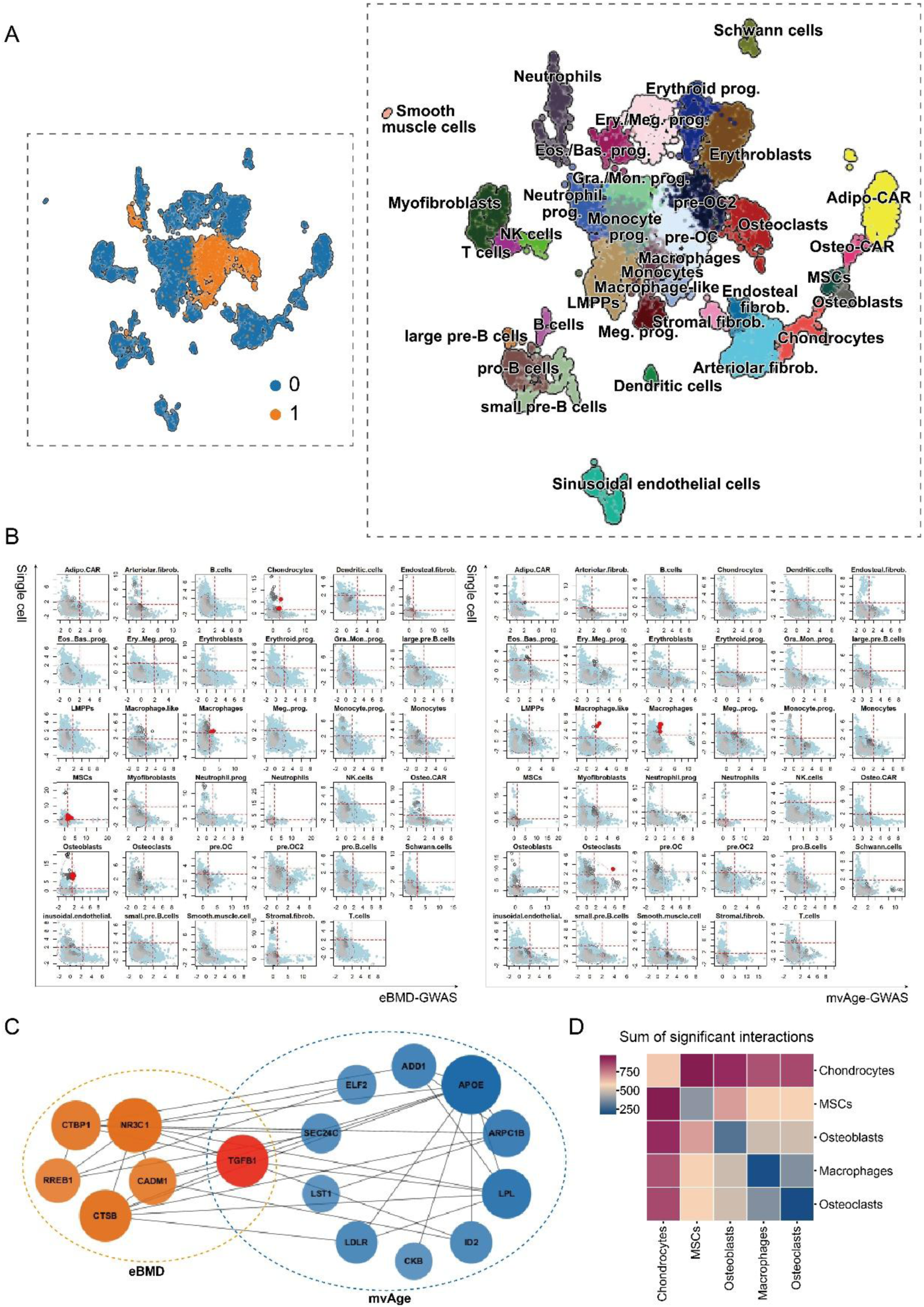
Integrative analysis of single-cell transcriptomics and GWAS summary statistics identifies macrophages as a pivotal cell type in bone aging. (A) UMAP projection of the integrated single-cell atlas. The left panel illustrates the batch integration of two distinct input datasets: dataset 0 (bone marrow niche) and dataset 1 (osteoclastogenesis dataset). The right panel displays the refined cell type annotations across the integrated atlas, covering a broad spectrum of bone marrow cell populations. Abbreviations: prog., progenitor; Meg., Megakaryocyte; Ery., Erythroid; OC., Osteoclasts; Gra., Granulocyte; Mon., Monocyte; Eos., Eosinophil; Bas., Basophil; Fibrob., Fibroblasts. (B) scGWAS analysis revealing cell type-specific enrichment for bone aging-related traits. Scatter plots show the enrichment of GWAS summary statistics for eBMD (left panel) and mvAge (right panel) across different cell types, with red dots indicating significant genetic enrichment. (C) Network diagram of key module genes identified through scGWAS. TGFB1 is highlighted in red as a central hub gene identified by both traits. (D) Significant ligand-receptor interactions between key cell types derived from scGWAS results.

We utilized the scGWAS approach (Jia et al. 2022) to integrate scRNA-seq and GWAS datasets related to osteoporosis (eBMD) and aging (mvAge). The results showed that MSCs, osteoblasts, chondrocytes, and macrophages were associated with eBMD; while osteoclasts and macrophages were associated with aging (Supplementary Table 2). The macrophages are involved in both processes (Figure 2B). The scGWAS analysis relies on modules constructed from protein-protein interaction networks. In the significant modules identified by the eBMD-scGWAS analysis, important osteoclast-regulating proteins such as cathepsin B (CTSB) and transforming growth factor beta 1 (TGFB1), were found (Figure 2C). The CTSB is a lysosomal protease highly expressed in activated macrophages and involved in osteoclastogenesis (Qiu et al. 2025; Li, Liang, et al. 2021); while the TGFB1 is a key cytokine regulating macrophage polarization and immunomodulation (Gong et al. 2012). The aging-related modules included important aging genes such as *APOE* and *TGFB1* (Figure 2C). The *APOE*, a known aging-associated gene, has been implicated in macrophage lipid metabolism and inflammatory responses (Montagne et al. 2020; Alzheimer’s Disease Neuroimaging Initiative et al. 2017; Phu et al. 2023). The presence of these macrophage-relevant genes and pathways in osteoporosis and aging-related modules supports the notion that macrophages are central mediators and *TGFB1* is a key gene in the bone aging process. To further elucidate the role of macrophages in bone aging, cell communication analysis was performed using CellPhoneDB (Garcia-Alonso et al. 2022). The results revealed extensive ligand-receptor interactions between macrophages and other cell types, particularly chondrocytes and mesenchymal stem cells (MSCs) (Figure 2D). Macrophages may regulate the osteogenic and chondrogenic differentiation of mesenchymal stem cells (MSCs), while MSCs and chondrocytes can, in turn, influence macrophage polarization (Zhang et al. 2020; Pajarinen et al. 2019).

Single-cell sequencing data derived from the human bone marrow niche were also utilized to perform scGWAS for validation in humans. The results identified macrophages as the cell type most strongly associated with mvAge, whereas osteoblasts showed the strongest correlation with eBMD (Supplementary Figure 4). Given that macrophages serve as the precursor cells for osteoclasts, which are central to bone remodeling, and play a critical role in human bone aging, these findings align with previous observations in mouse models. Collectively, these results suggest that macrophages may play a pivotal regulatory role in linking the aging process to osteoporosis.

### Macrophage remodeling, epigenetic alterations and perturbed cell communication in aging

To investigate the age-related changes in the number and proportion of bone marrow macrophages, we analyzed two independent scRNA-seq datasets derived from the bone marrow of mice at different ages (The Tabula Muris Consortium et al. 2018; Ma et al. 2022). Although the majority of bone marrow cells were of immune origin, macrophages constituted only a small fraction of the total cell population (Figure 3A). Notably, both datasets revealed a consistent and substantial decline in the proportion of bone marrow macrophages with advancing age (Figure 3B, Supplementary Figure 5), a trend similar to the changes observed in skeletal muscle microenvironment (Kedlian et al. 2024). Further re-clustering of macrophages identified five distinct subpopulations: IFN-stimulated, proliferating, MHC-II^+^, lipid-associated, and inflammatory macrophages (Supplementary Figure 6). Compositional analysis showed that the proportion of lipid-associated macrophages increased with age within the total macrophage pool, whereas the IFN-stimulated and inflammatory subsets decreased. In contrast, the proportion of MHC-II^+^ macrophages exhibited an increase during late life (months 18–24) (Figure 3C). Furthermore, scGWAS analysis suggested that MHC-II^+^ macrophages are associated with both eBMD and mvAge, indicating their potential involvement in bone aging processes (Supplementary Table 7).

**Figure 3.**
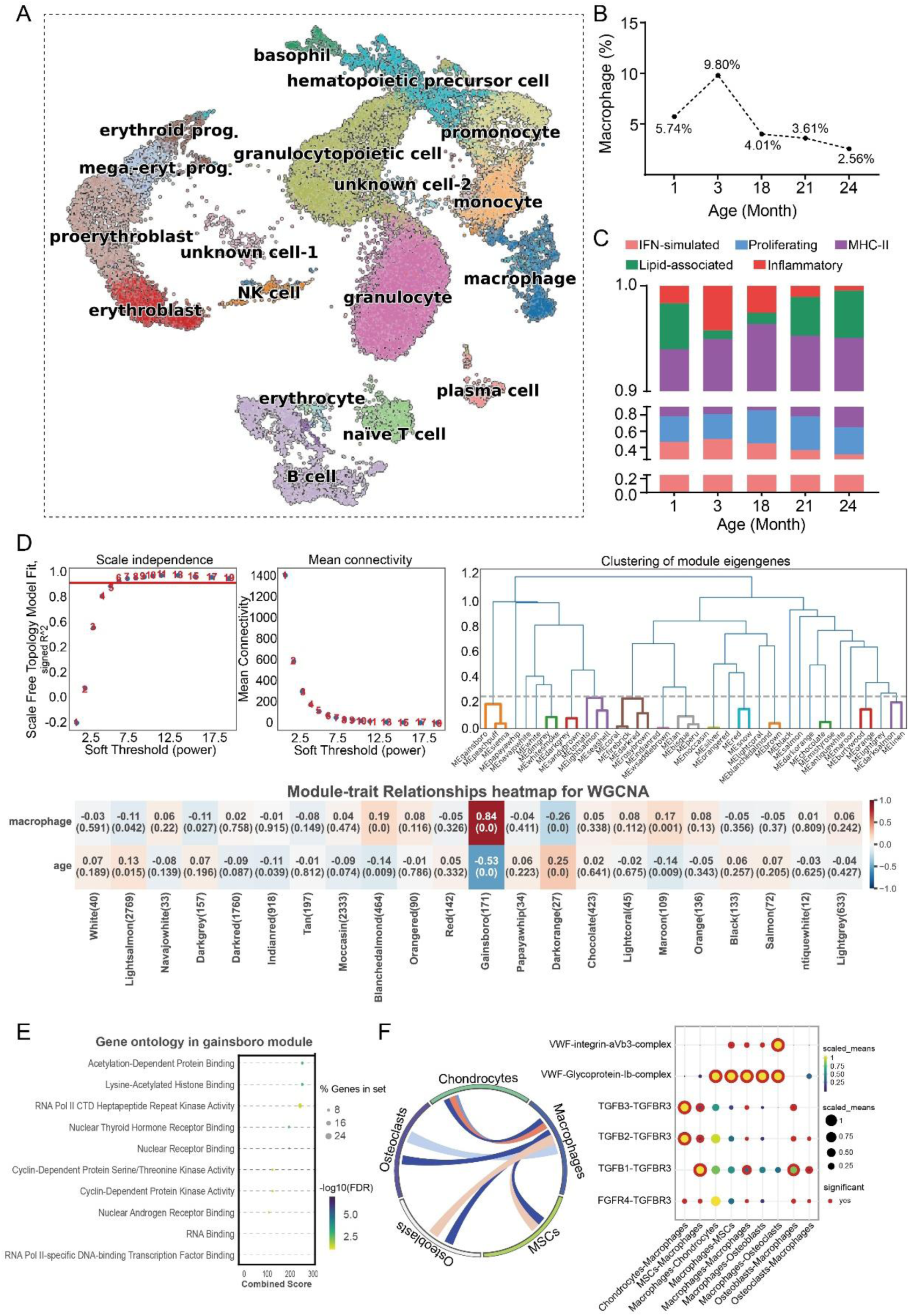
TGFB signaling mediates the age-related decline of macrophages. (A) UMAP visualization of the single-cell transcriptomic landscape of mouse bone marrow across different ages. (B) Quantification showing that the proportion of macrophages decreases with age. (C) Stacked bar plot illustrating the proportional changes of macrophage subclusters (e.g., IFN-simulated, Proliferating, MHC-II, Lipid-associated, Inflammatory) across age groups. (D) scWGCNA analysis identifying the “Gainsboro” module associated with macrophage abundance. Top panels show scale-free topology fit and mean connectivity for soft-thresholding selection, and clustering of module eigengenes. The bottom heatmap displays module-trait relationships, highlighting the Gainsboro module’s positive correlation with macrophages and negative correlation with age. (E) Gene Ontology (GO) enrichment analysis of genes within the Gainsboro module. (F) Cell-cell communication analysis in the context of bone aging. Genes from the Gainsboro module were used as the input gene set to reveal specific ligand-receptor interactions, highlighting TGFB signaling pathways between macrophages and other cell types.

To identify key gene networks associated with macrophages during aging, we performed a single-cell weighted gene co-expression network analysis (scWGCNA) (Morabito et al. 2023; Rezaie et al. 2023). First, metacell transcriptomic profiles were generated from the scRNA-seq datasets to reduce technical noise and enhance biological signal. The optimal soft-thresholding power for network construction was determined by fitting the network to a scale-free topology, with the threshold selected based on the highest R^2^ value that approached the scale-free topology model fit (Figure 3D). Using hierarchical clustering with the dynamic tree cut method, we identified 22 distinct gene co-expression modules (Figure 3D). Among these, the Gainsboro module, consisting of 171 genes, exhibited the strongest positive correlation with the macrophage proportion and negative correlation with age (Figure 3D, Supplementary Table 8). To explore the functional implications of this module, we conducted Gene Ontology (GO) enrichment analysis, which revealed that Gainsboro module genes were predominantly involved in various regulatory processes, including transcriptional regulation, chromatin remodeling, and cellular signaling. Notably, epigenetic regulatory processes emerged as the most significantly enriched category (Figure 3E). Within this category, BRD2, PSME4, BAZ2A, and ZZEF1 were specifically associated with lysine-acetylated histones and acetylation-dependent protein binding (Supplementary Table 9), suggesting a potential link between age-related changes in macrophages and alterations in epigenetic mechanisms within the bone marrow microenvironment.

By treating the genes within the Gainsboro module as differentially expressed genes (DEGs), we performed cell communication analysis again. The TGF-β pathway and von Willebrand Factor (VWF) complex emerged as potential interactions (Figure 3F). These findings suggest that TGF-β and VWF may serve as key molecular mediators through which macrophages contribute to age-related signaling in the bone marrow microenvironment.

### Shared genetic architecture of BMD and aging

Both scWGCNA and Cellphonedb rely on known protein-protein interactions. To identify novel key bone aging genes, we employed shared genetic methods. A leftward deflected from the expected null line in the stratified conditional Q-Q plot indicates SNP enrichment for the eBMD as a function of the association with mvAge (Figure 4A). The conjunctional false discovery rate (conjFDR) analysis identified approximately 160 SNPs that were significantly associated with both phenotypes simultaneously (Supplementary Figure 7, Supplementary Table 10). To further validate these findings, a local genetic correlation method (Zhang et al. 2021) was applied (Supplementary Table 5), and chromosomal regions harboring these 20 SNPs showed polygenic overlap between eBMD and mvAge (Figure 4B, Supplementary Table 5). eQTL mapping linked these 20 SNPs to 43 genes; notably, *HNRNPUL1* (Heterogeneous Nuclear Ribonucleoprotein U Like 1) was the sole gene overlapping with the “Gainsboro” module identified by WGCNA (Figure 4C). The ccFDR-significant SNP rs12975920 resides within the *HNRNPUL1* gene body (Figure 4D). Notably, *HNRNPUL1* is located in close physical proximity to *TGFB1* on the chromosome (Figure 4D). These findings suggest that *HNRNPUL1* may be a novel gene involved in both aging and osteoporosis at the same time.

**Figure 4.**
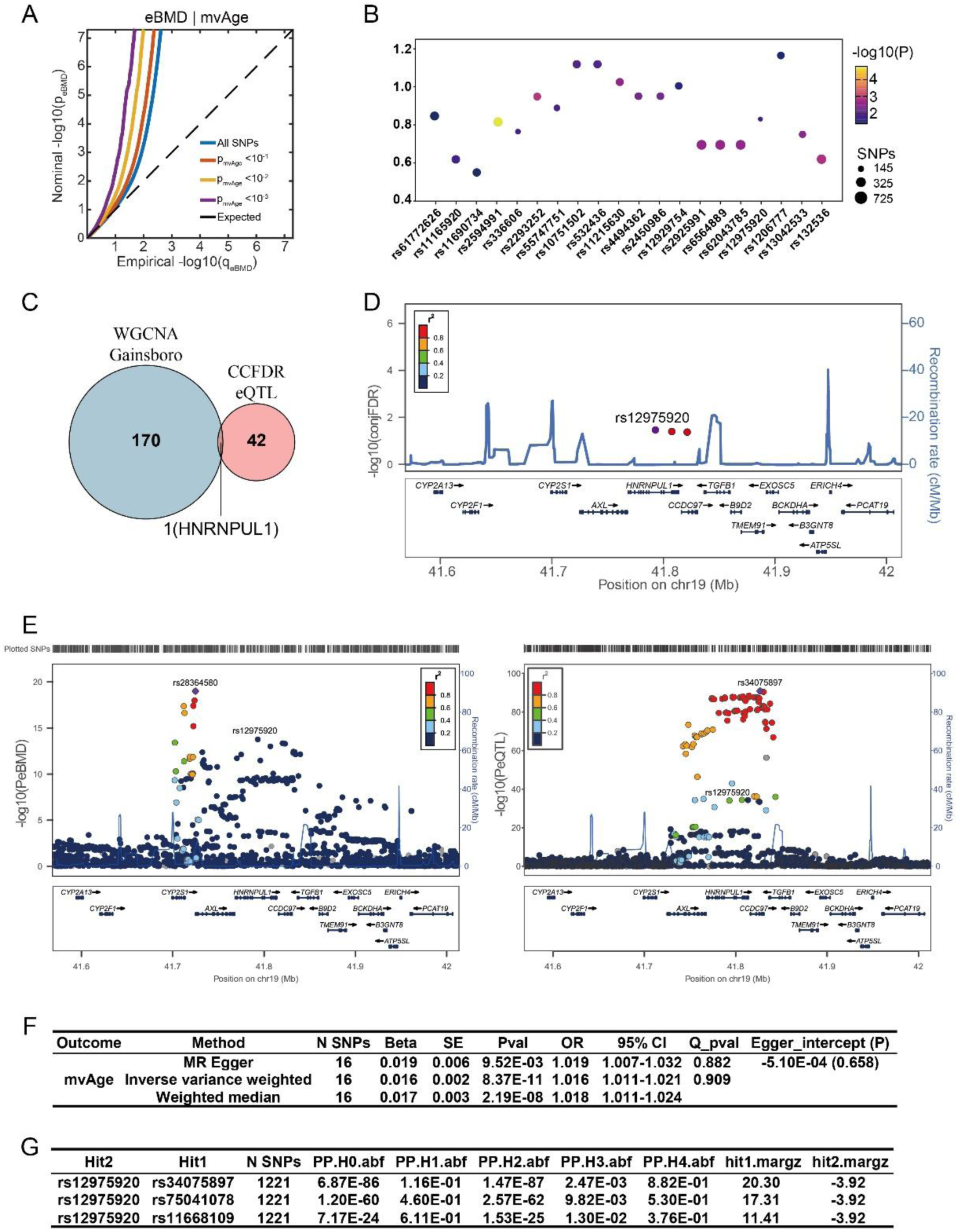
Shared genetic methods identified *HNRNPUL1* as an important gene in bone aging. (A) Conditional quantile-quantile (QQ) plots of eBMD associations stratified by the significance of associations with mvAge at p<0.1, p<0.01, p<0.001. (B) Local genetic correlation analysis of genomic regions containing the corresponding SNPs. (C) Venn diagram illustrating *HNRNPUL1* as the only overlapping gene identified by WGCNA (Gainsboro) and those mapped by ccFDR/eQTL analysis. (D) Regional association plot (LocusZoom) showing the ccFDR-significant SNP rs12975920 located within the *HNRNPUL1* gene body. The blue line indicates the recombination rate. (E) Regional association plots for eBMD at the *HNRNPUL1* locus. Both SNPs located within the *HNRNPUL1* gene body and eQTL-related SNPs demonstrate significant associations with eBMD. (F) MR analysis reveals genetically determined *HNRNPUL1* is associated with mvAge. (G) Colocalization analysis of eQTL and mvAge GWAS signals, displaying posterior probabilities for shared causal variants (PP.H4 > 0.8).

The SNPs located within the gene body of *HNRNPUL1*, or those that are significantly associated with its expression levels based on expression quantitative trait locus (eQTL) analyses, show strong association with eBMD (Figure 4E). This finding highlights a potential functional link between *HNRNPUL1* and the pathogenesis of osteoporosis. Selecting SNPs significantly related to *HNRNPUL1* expression using eQTL as instrumental variables, and conducting summary-level statistics on mvAge as outcome, we performed Mendelian Randomization (MR) to assess the role of *HNRNPUL1* in aging. The MR results provide evidence supporting a causal relationship *HNRNPUL1* expression and aging-related phenotypes (Figure 4F). Moreover, colocalization analysis indicates that this observed association is mediated through the regulation of *HNRNPUL1* gene expression (Figure 4G). Taken together, these results suggest that *HNRNPUL1* may play a role in bone aging.

### A potential role for *TGFB1* and *HNRNPUL1* in macrophage function and osteoclast differetiation

Macrophages serve as the precursor cells of osteoclasts. To elucidate the specific mechanisms of the *TGFB1* and *HNRNPUL1* gene in osteoporosis, we first determine whether *TGFB1* and *HNRNPUL1* influence osteoclast differentiation. From the integrated scRNA-seq atlas, we selected cell types pertinent to osteoclast differentiation. We calculated an osteoclast differentiation score based on the expression of canonical markers (*ACP5*, *MMP9*, and *CTSK*). This score increased progressively during differentiation, validating the reliability of our atlas for tracking the osteoclast lineage. Conversely, the expression levels of *TGFB1* and *HNRNPUL1* decreased during macrophage differentiation, suggesting that both genes may regulate macrophage fate decisions, thereby impacting osteoclastogenesis (Figure 5A).

**Figure 5.**
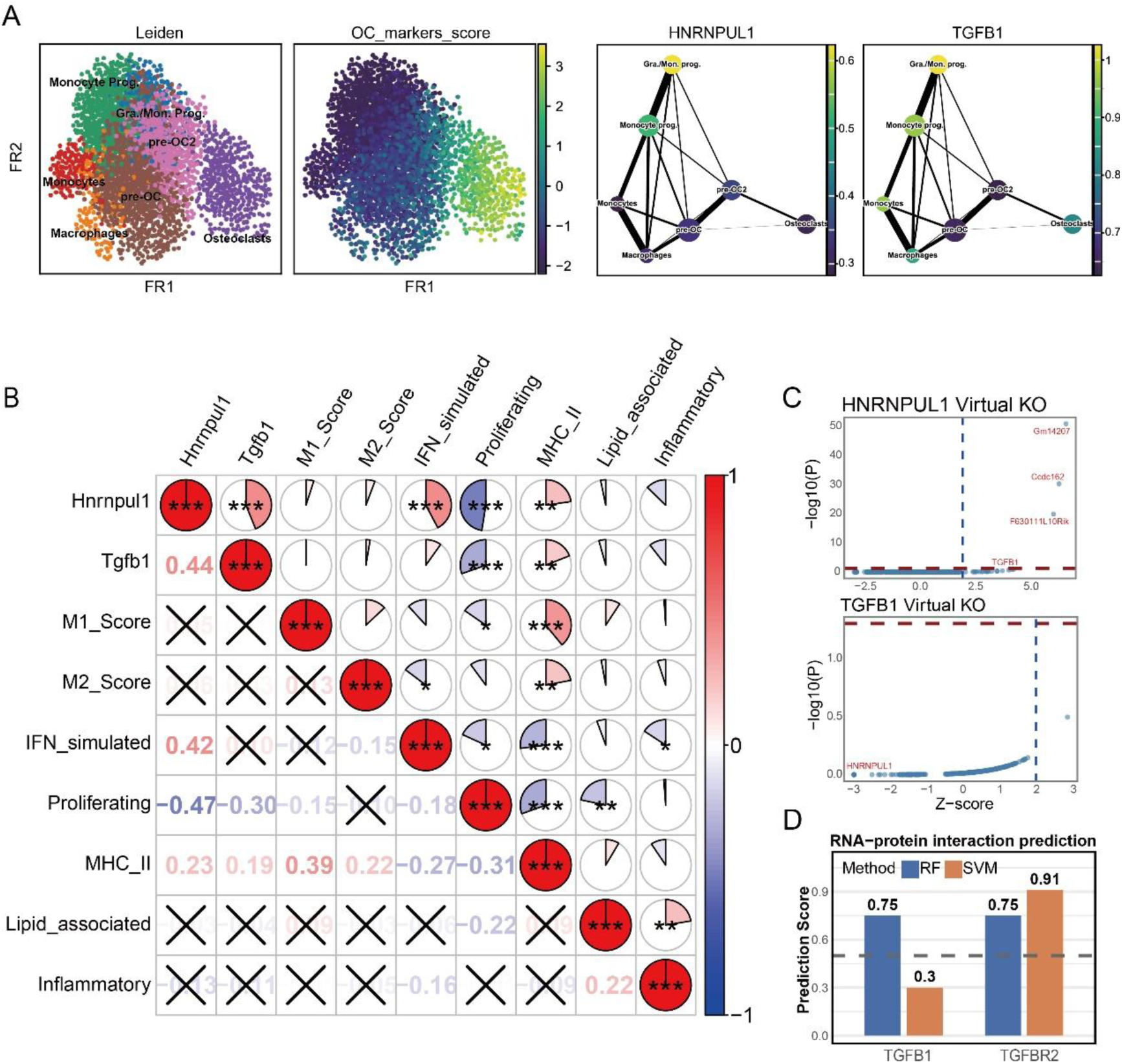
Expression dynamics of *HNRNPUL1* and *TGFB1* during osteoclast differentiation and their association with specific macrophage subclusters. (A) Trajectory plots illustrating the expression levels of *HNRNPUL1* and *TGFB1*, which decrease during macrophage differentiation. (B) Correlation matrix displaying relationships between gene expressions (*HNRNPUL1*, *TGFB1*), classical polarization scores (M1, M2), and specific macrophage subcluster scores (*: *P* < 0.01, **: *P* < 0.01, ***: *P* < 0.001). (C) Knockout of *HNRNPUL1* (left) or *TGFB1* (right) did not result in significant changes in the expression of the other gene, indicating a lack of direct transcriptional regulation between them. (D) Bar chart showing computational prediction scores (using RF and SVM methods) for RNA-protein interactions. A score > 0.5 suggests a high likelihood of binding.

Macrophages undergo activation and polarization into functionally distinct subpopulations by specific simulation from microenvironment. M2 polarization macrophages can promote pathologic angiogenesis as well as organ fibrosis, and secrete TGFB1 to stimulate osteogenic differentiation (Cai, Lu, et al. 2023; Wang et al. 2022; He et al. 2021; Oshi et al. 2020). Consequently, we investigated whether HNRNPUL1 also influences macrophage polarization. We compiled marker genes for M1 polarization (*NOS2*, *CD86*, *CXCL10*, *IL1B*, *CXCL9*) and M2 polarization (*ARG1*, *MRC1*, *CD163*, *RETNLA*, *CHIL3*, *CD36*) (He et al. 2021; Lawrence & Natoli 2011; Martinez et al. 2006). After calculating polarization score using ‘tl.score_genes’ function from Scanpy (Wolf et al. 2018), we observed no significant correlation between *HNRNPUL1* or *TGFB1* expression and classical M1/M2 polarization states (Figure 5B). Furthermore, no distinct macrophage subclusters exhibited exclusive M1 or M2 features (Supplementary Figure 8A). However, when scoring cells based on markers defining specific macrophage subclusters, both *HNRNPUL1* and *TGFB1* showed strong associations with the “Proliferating” and “MHC-II^+^” macrophage subclusters (Figure 5B; Supplementary Figure 8B). Notably, the expression levels of markers for Proliferating and MHC-II^+^ macrophages did not show distinct correlations with age (Supplementary Figure 8C).

To further infer the regulatory relationship between *HNRNPUL1* and *TGFB1*, we performed virtual knockout analyses. The results indicated that neither gene significantly influenced the expression level of the other (Figure 5C). Given that *HNRNPUL1* encodes an RNA-binding protein, we predicted its interaction with *TGFBs* mRNA, the computational predictions suggested a high potential for binding (score > 0.5), a finding corroborated by data from the CLIP database (Figure 5D; Supplementary Table 12). Collectively, these results suggest that *HNRNPUL1* may regulate *TGFBs* RNA splicing, suggesting an *HNRNPUL1-TGFB* signaling axis that influences macrophage cell fate and function.

## Discussion

With the increasing prevalence of population aging, understanding the specific mechanisms of bone aging has become crucial. Our research highlights there is a bilateral causal relationship between systematic aging and osteoporosis. To date, many studies offer partial or indirect evidence supporting this bilateral causality. The gradual loss of bone with advancing age indicates that aging can induce osteoporosis (Curtis et al. 2015). Although the impact of osteoporosis on aging is less studied, evidence suggests that conditional deletion of osteocytes in a mouse model results in not only bone mass loss but also a senescence-associated secretory phenotype (SASP) and shortened lifespan, indicating that osteoporosis also accelerates aging (Ding et al. 2022).

In this study, we corroborated this bidirectional causality using regression analyses of individual phenotypic data and MR analyses of GWAS summary statistics. Therefore, the prevention and treatment of osteoporosis may offer a promising strategy for delaying systemic aging, underscoring the urgent need for more detailed research into the cell types and genes that link aging and osteoporosis.

We found macrophages play a critical role in bone aging and may serve as a communication bridge between systemic aging and osteoporosis, with *TGFB1* and *HNRNPUL1* emerging as two critical regulators. As key components of the innate immune system, macrophages are central to identifying and eliminating pathogens, regulating inflammatory responses, and promoting tissue repair. However, with advancing age, individuals develop a persistent proinflammatory state, and macrophage function gradually deteriorates, increasing susceptibility to infections and chronic inflammation (Gibon et al. 2017). Macrophages are heterogeneous, while classic models categorize them into classically activated (M1) or alternatively activated (M2) subsets (Martinez et al. 2006; Cui et al. 2019), increasing evidence suggests this binary model fails to capture the full complexity of macrophage heterogeneity and function. We classified macrophages into five distinct clusters: IFN-stimulated, proliferating, MHC-II^+^, lipid-associated, and inflammatory. Notably, MHC-II^+^ macrophages showed a significant association with bone aging, as did the expression of *HNRNPUL1* and *TGFB1*. This finding aligns with reports that a significant enrichment of MHC-II+ macrophages in high-risk thin-cap fibroatheromas (TCFAs) relative to stable plaques. (Xie et al. 2026). In TCFAs, these antigen-presenting cells fuel plaque progression by activating local T-cell inflammatory cascades. The parallel accumulation of this specific macrophage subset in both aging bone and vulnerable plaques suggests that targeting antigen-presenting macrophages could offer a promising dual-purpose therapeutic avenue for treating bone aging and preventing acute cardiovascular events. *TGFB1* and *HNRNPUL1*, two adjacent genes on chromosome 19 exhibiting coordinated age-related expression, may jointly regulate bone aging through macrophage-mediated mechanisms. *TGFB1*, a major cytokine in bone matrix, is well established in osteoporosis research for its role in promoting bone remodeling, particularly by simulating migration of bone marrow stromal cells (BMSCs) (Tang et al. 2009; Chen et al. 2012). Beyond bone, TGF-β signaling is increasingly linked to systemic aging, contributing to chronic inflammation and fibrosis (Peng et al. 2025; Farhat et al. 2025), although its direct role in bridging aging and osteoporosis remains underexplored. Similarly, *HNRNPUL1*, an RNA-binding protein involved in DNA damage response via NBS1 recruitment (Gurunathan et al. 2015; Polo et al. 2012), shows causal effects on aging based on our Mendelian randomization and colocalization analyses. Zebrafish *Hnrnpul1* mutants exhibit skeletal defects, suggesting a potential role in both bone development and osteoporosis (Blackwell et al. 2022). Functioning as a splicing factor, *HNRNPUL1* is predicted to bind *TGFB* mRNA, forming an *HNRNPUL1*-*TGFB1* axis that regulates macrophage function and bone aging.

Several limitations exist in this study. First, integrating GWAS summary statistics with single-cell sequencing data using methods such as sc-eQTL analysis would strengthen the validity of our observations. While scGWAS relies on average expression within cell populations, sc-eQTL utilizes expression data from individual cells; however, publicly available sc-eQTL data for bone tissue remain scarce. Additionally, the limited number of macrophages captured constrained the ability to perform deeper subclustering, potentially limiting the resolution of macrophage subset characterization. Finally, the functional role of the *HNRNPUL1*-*TGFB1* axis requires further experimental validation and exploration.

In summary, this study presented genetic proof of a two-way causal connection between aging and osteoporosis. It also revealed a macrophage-centered mechanism governed by *TGFB1* and *HNRNPUL1*. Collectively, these findings offer fresh perspectives and potential therapeutic targets for age-associated bone loss.

## Data Availability

No specific data prodeced

https://github.com/tinker710/bone_aging

## Author contributions

**Xin Li**: Formal analysis, Visualization, Writing-Original Draft; **Meng-Yuan Yang**: Validation, Writing-Original Draft; **Si-Rui Gai**: Investigation, Software; **Peng Wei**: Investigation; **Zeng-Hui Gu**: Investigation; **Yue-Zhou Wu**: Investigation; **Ming-Yu Han**: Software, Methodology; **Jia-Sheng Yu**: Investigation; **Wang-Jun Chen**: Investigation; **Zhen-Rui Liao**: Investigation; **Jia-Xuan Gu**: Software, **Jia-Dong Zhong**: Visualization; **Pian-Pian Zhao**: Resources; **Ke Zhu**: Resources, **Ching-Lung Cheung**: Conceptualization; **David Karasik**: Conceptualization; **Hou-Feng Zheng**: Conceptualization, Writing-Review and Editing, Supervision, Funding acquisition.

## Acknowledgements

This work was supported by the National Natural Science Foundation of China (#82673195), by the National Key R&D Programme of China (#2024YFC3405703), and by the National Natural Science Foundation of China (#82370887). We would like to thank the UK Biobank and GTEx Consortium. This work was supported by Beijing GuoKe Biotechnology Co., LTD (Beijing, China).

## Data Availability Statement

The data that supports the findings of this study are openly available and detailed information is in the supplementary material of this article.

---

**Supplementary Figure 1.**
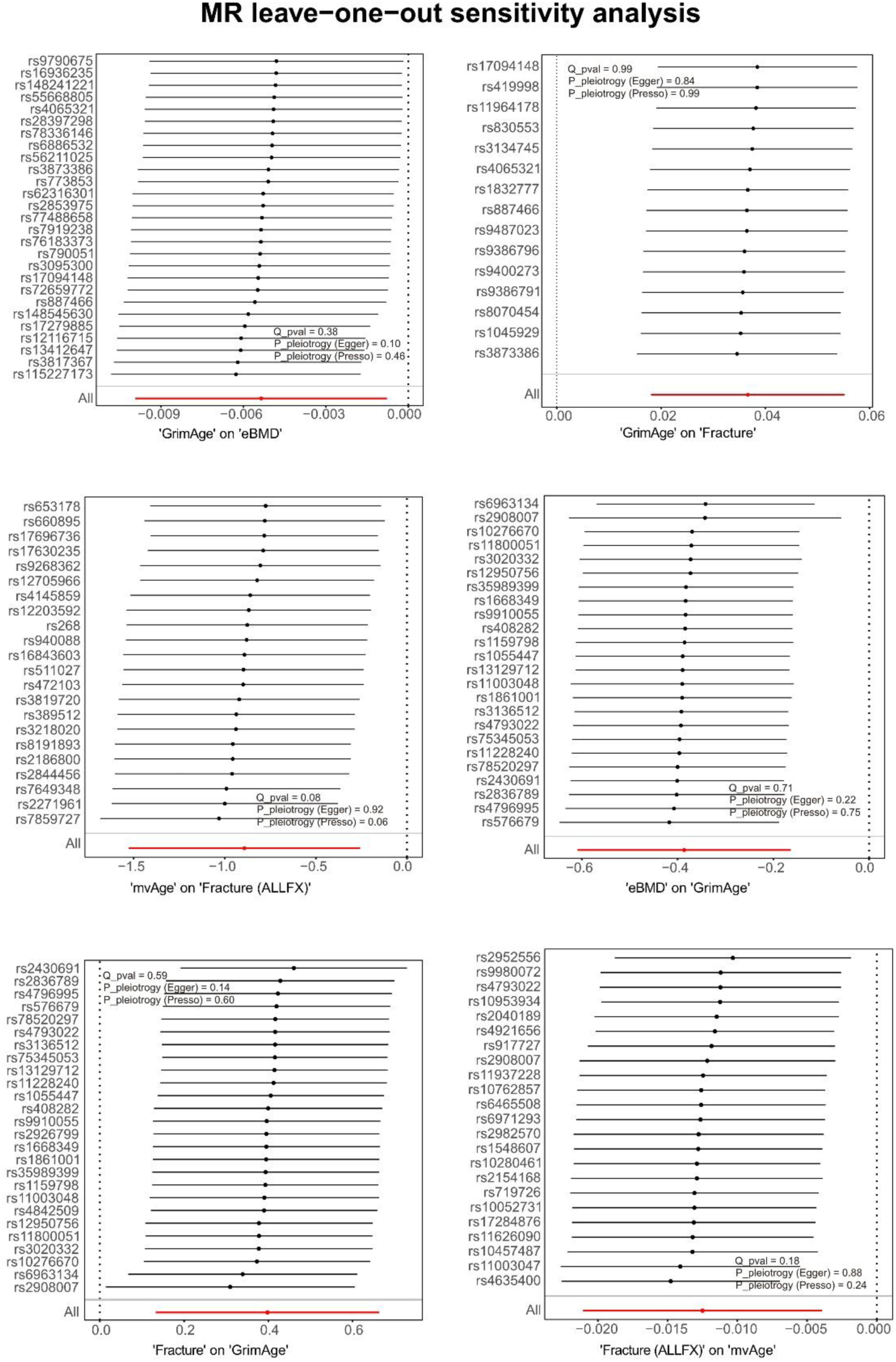
Sensitivity analyses to evaluate the robustness of the MR results. Q_pval is the pvalue of Cochran’s Q test. Q_pval < 0.05 suggest heterogeneity. MR-Egger regression and MR-PRESSO are used to identify horizontal pleiotropy. If P_pleiotrogy (Egger) and P_pleiotrogy (Presso) > 0.05, there is no evidence of horizontal pleiotropy.

**Supplementary Figure 2.**
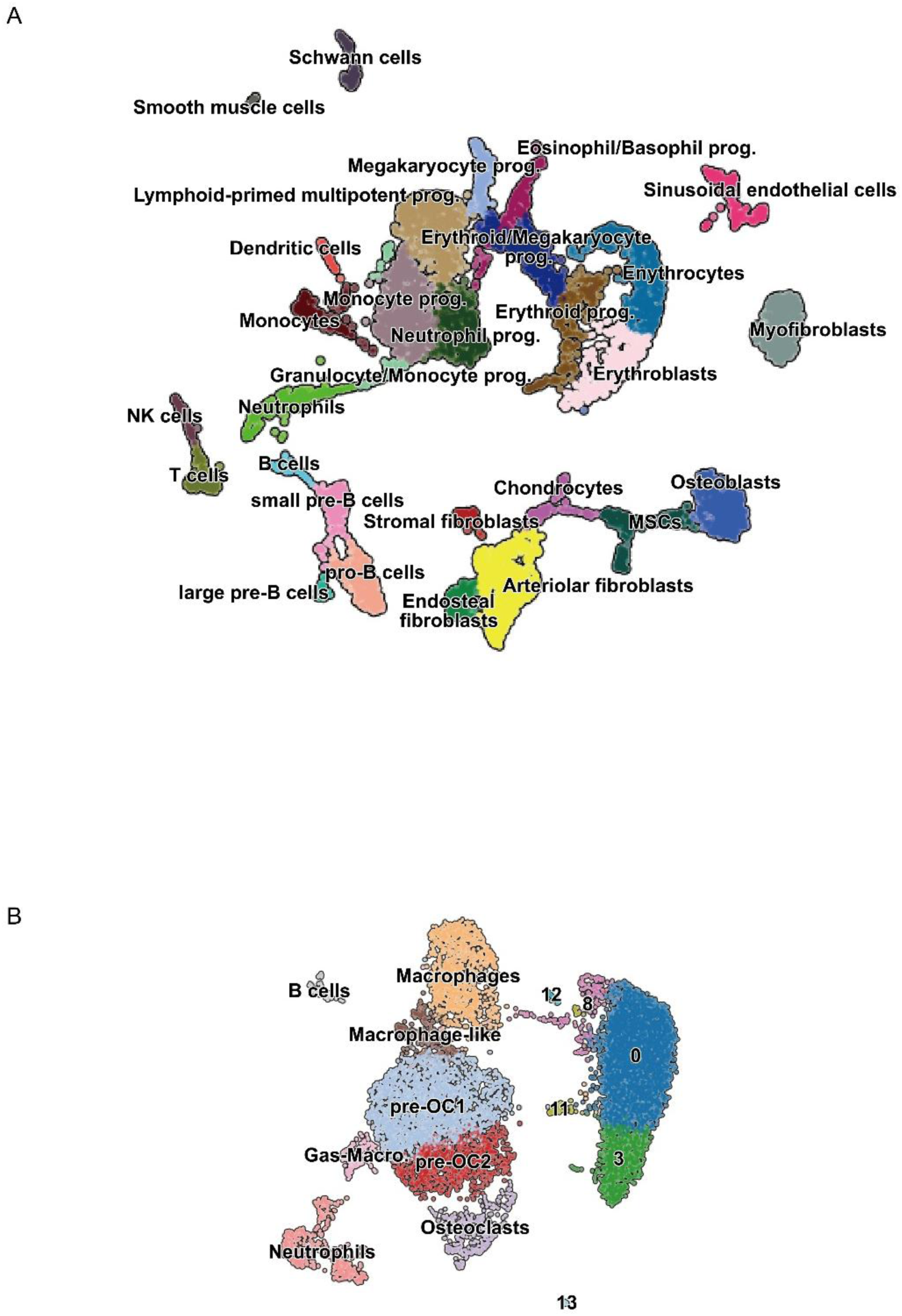
UMAPs of bone remodeling cell types. (A) UMAP of cell types related to immune cells, MSCs, osteoblasts differentiated from MSCs and others. (B) UMAP of cell types related to osteoclastogenesis.

**Supplementary Figure 3.**
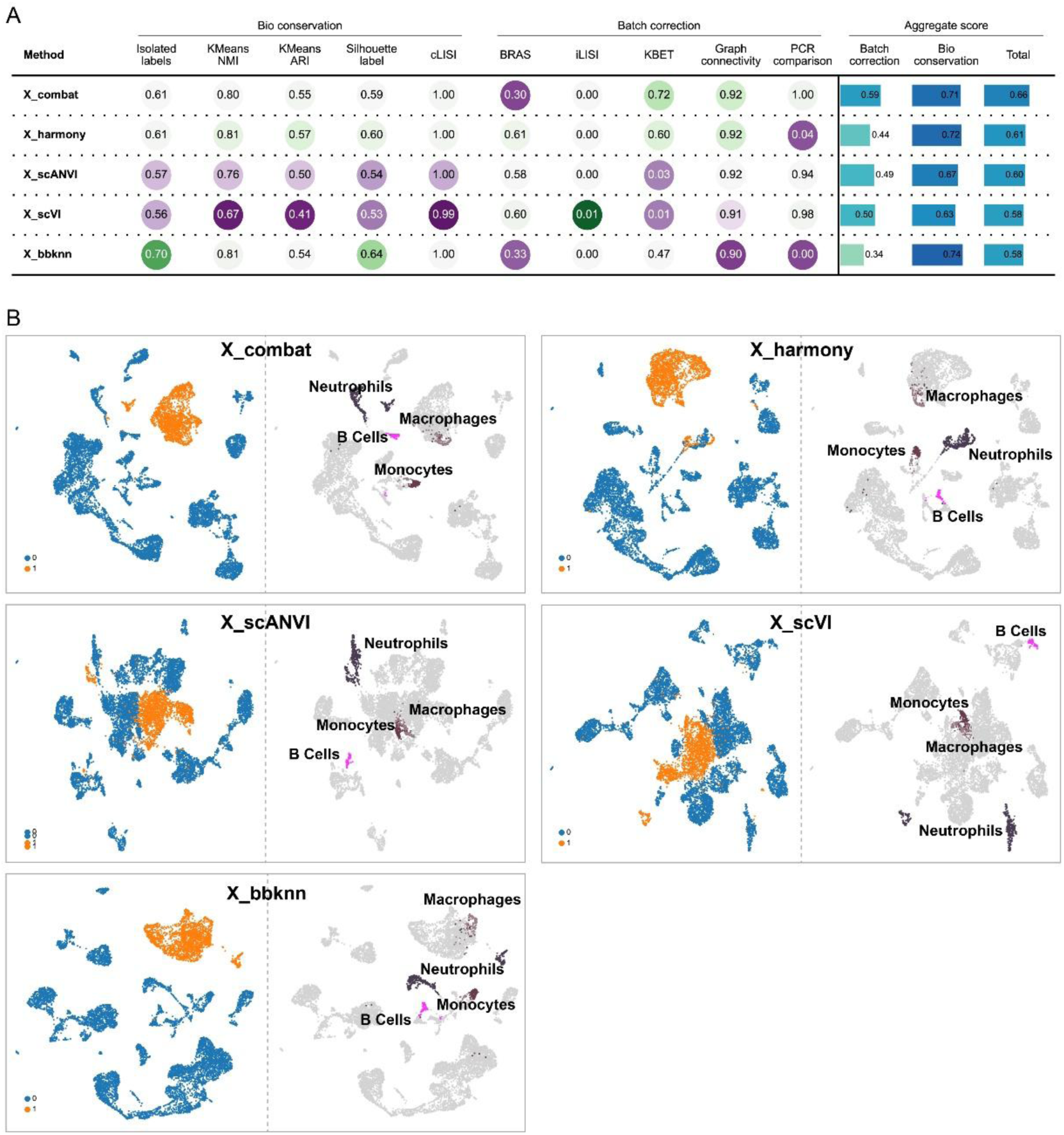
Benchmarking of integration using different methods on single-cell sequencing data. (A) Quantitative evaluation of 5 integration methods (Combat, Harmony, scANVI, scVI and BBKNN) across three categories: Biological conservation, Batch correction and Aggregate scores. (A) UMAP of integrated embeddings generated by each method. For each panel, left plot shows cell clustering colored by batch origin, while right plot shows shared cell types to assess biological structure preservation.

**Supplementary Figure 4.**
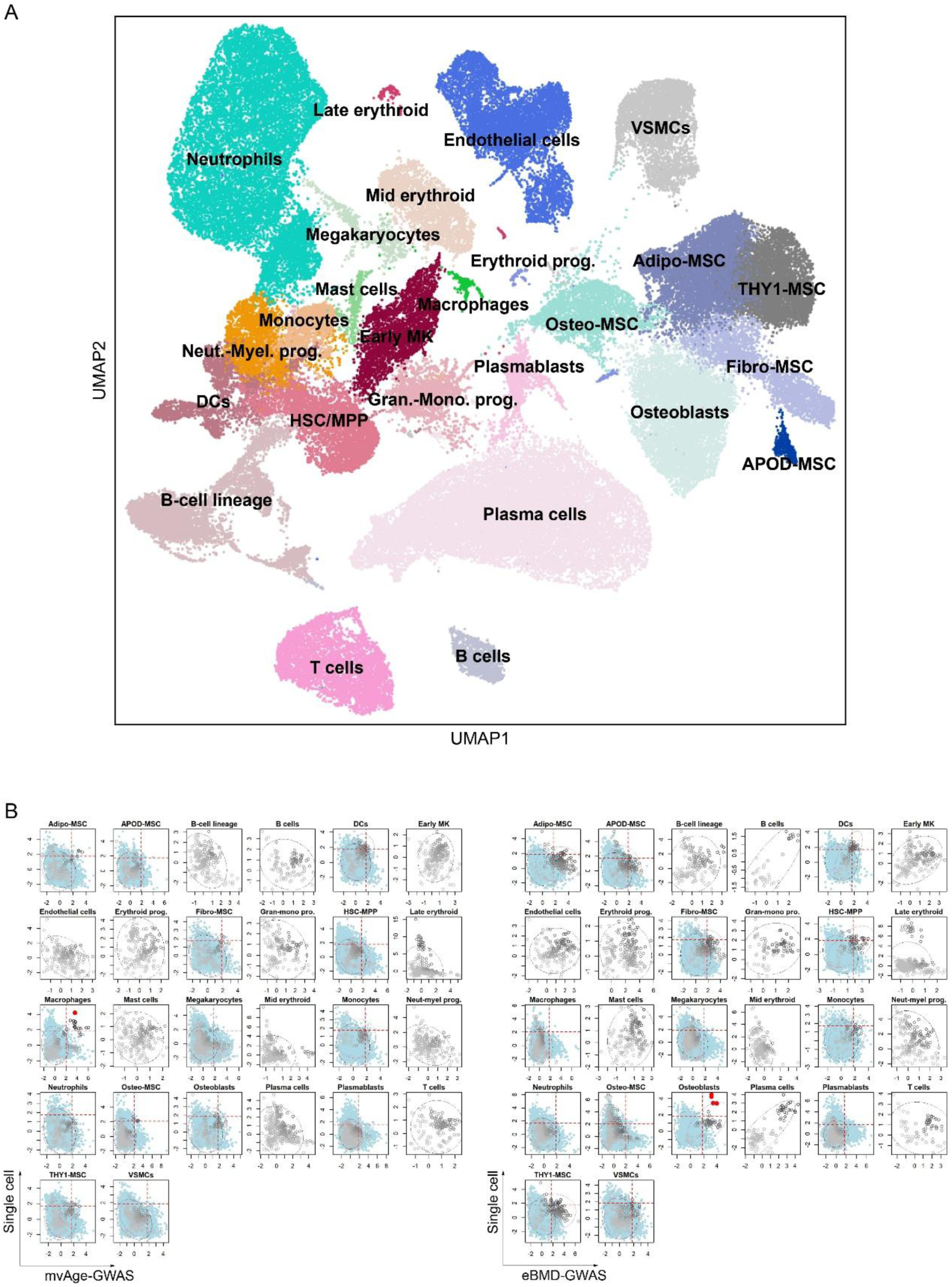
Integrative analysis of human bone marrow niche single-cell transcriptomics and GWAS summary statistics as a human validation. (A) UMAP visualization of annotated cell types of human bone marrow niche. (B) scGWAS analysis revealing cell type-specific enrichment for bone aging-related traits. Scatter plots show the enrichment of GWAS summary statistics for mvAge (left panel) and eBMD (right panel) across different cell types, with red dots indicating significant genetic enrichment.

**Supplementary Figure 5.**
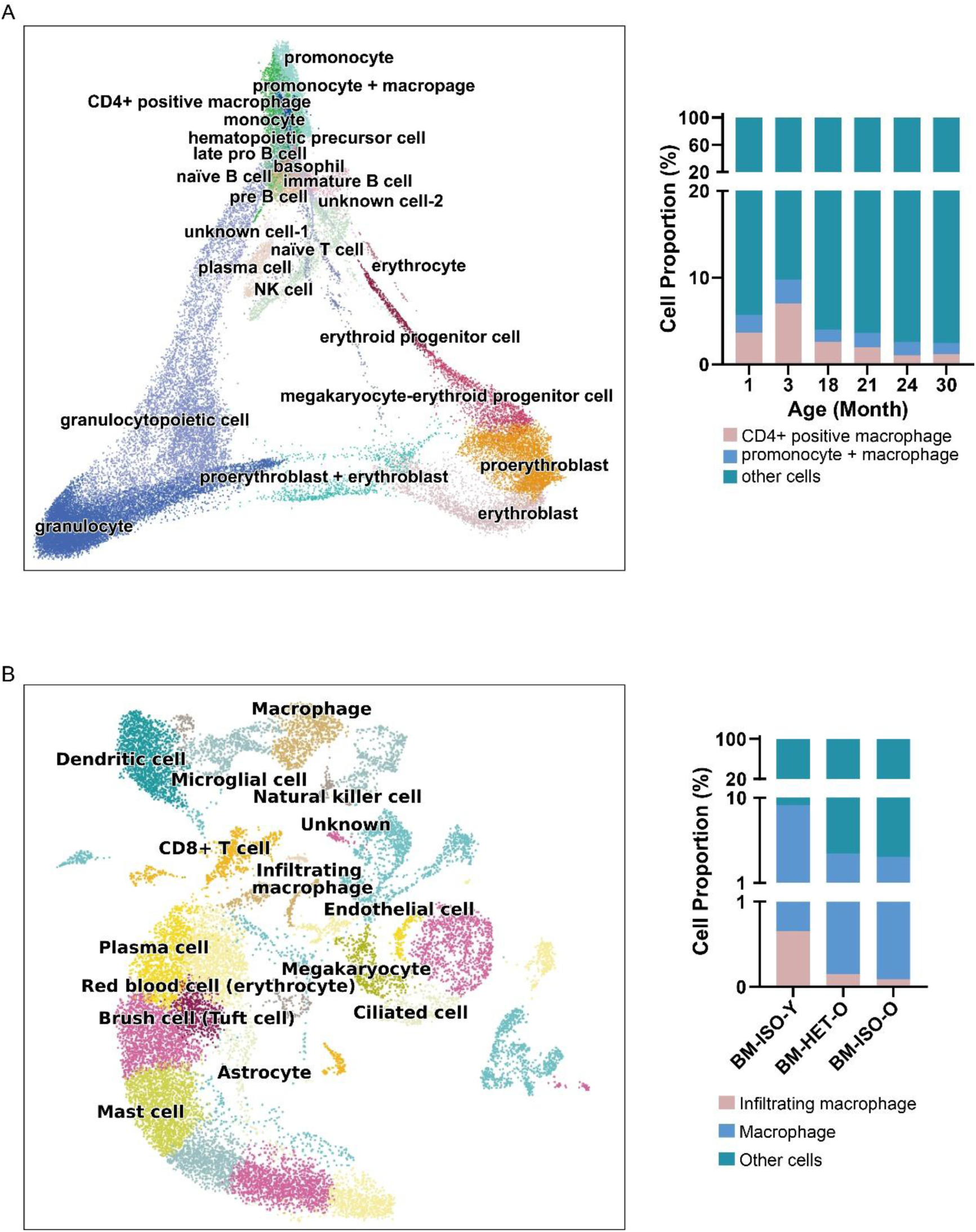
Marrow macrophage percentages at different ages. (A) Macrophage percentages across different ages, calculated with the annotation information provided in the original study. (B) Macrophage percentages in heterochronic parabiosis mice at different ages.

**Supplementary Figure 6.**
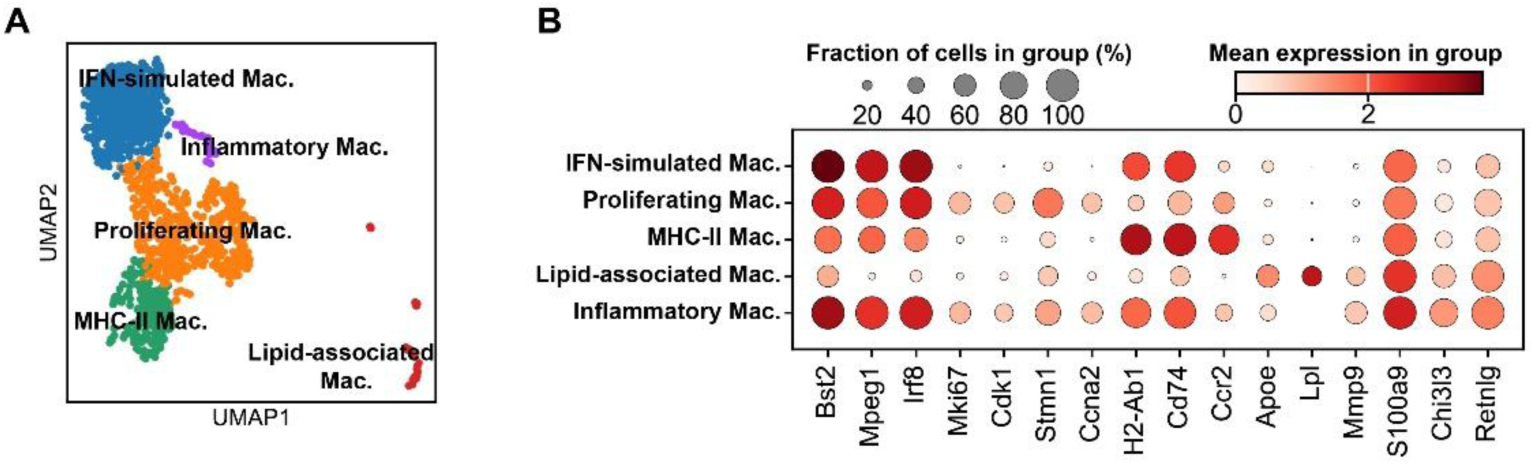
Characterization of macrophage subpopulations in mouse bone marrow niche. (A) UMAP visualization of macrophage heterogeneity, revealing five distinct clusters: IFN-simulated Macrophages, Inflammatory Macrophages, Proliferating Macrophages, MHC-II Macrophages and Lipid-associated Macrophages; (B) Dot plot illustrating the expression profiles of canonical marker genes across the identified macrophage subpopulations.

**Supplementary Figure 7.**
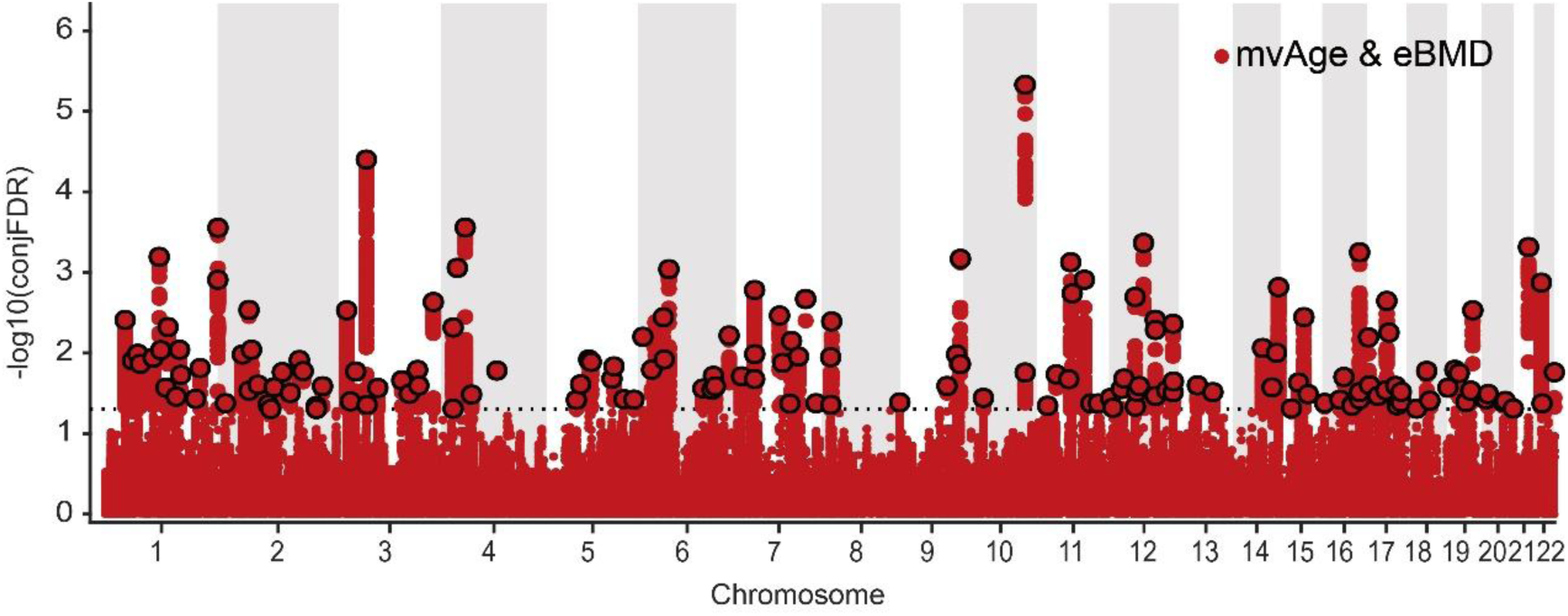
Shared genetic tools identify pleiotropic loci between mvAge and eBMD.

**Supplementary Figure 8.**
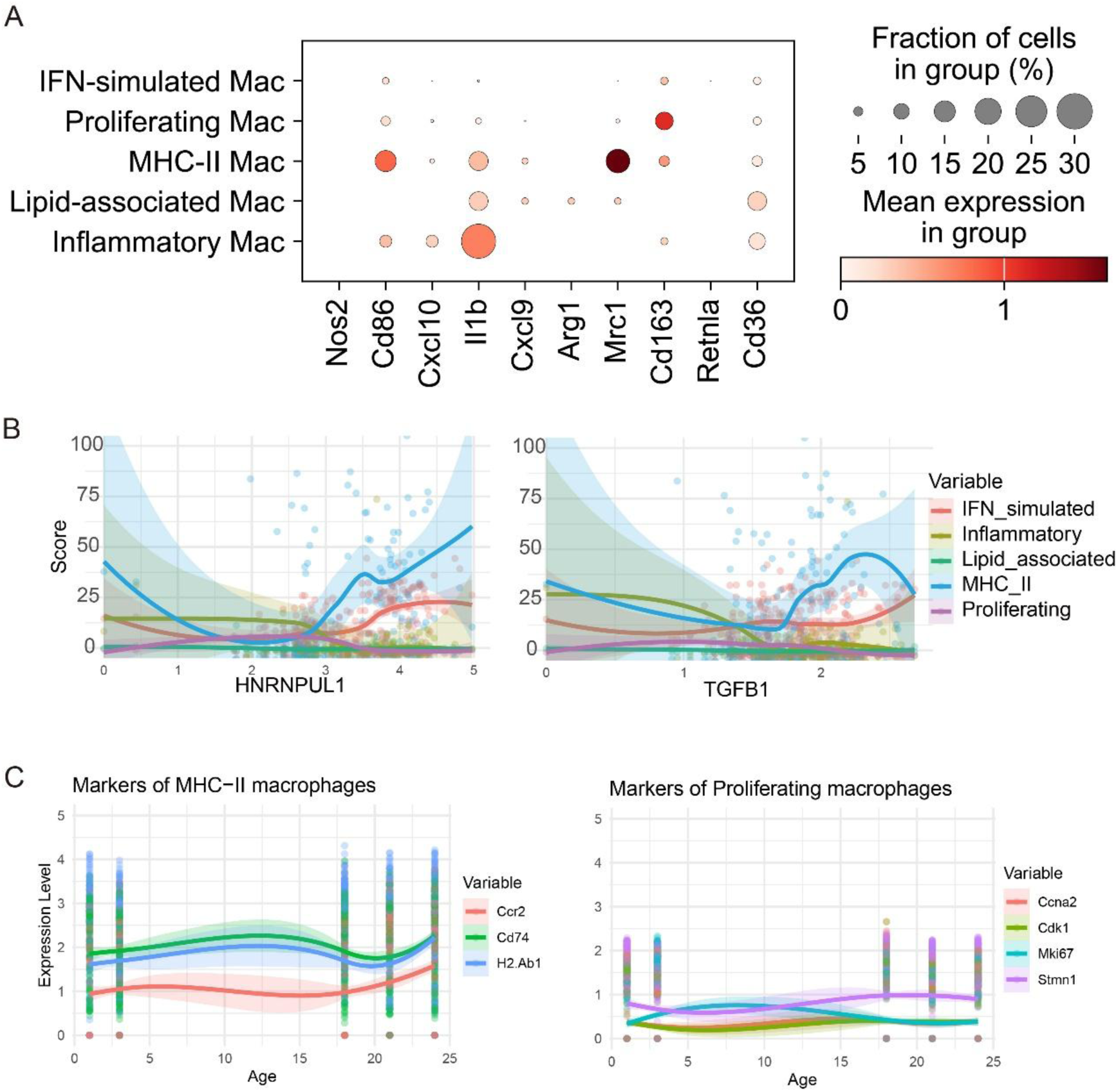
Characterization of macrophage subpopulations and their dynamics across age. (A) Dot plot illustrating the expression profiles of canonical marker genes of M1/M2 polarization across the identified macrophage subpopulations. (B) Line plots visualizing the positive correlation between *HNRNPUL1* or *TGFB1* expression levels and the scores for “Proliferating” and “MHC_II” macrophage subclusters. (C) Ridge plots and scatter plots depicting the expression trajectories of specific marker genes as a function of Age. The left panel highlights markers for MHC-II macrophages (*Ccr2*, *Cd74*, *H2-Ab1*), while the right panel shows markers for Proliferating macrophages (*Ccna2*, *Cdk1*, *Mki67*, *Stmn1*).

## Legends for supplementary tables

Supplementary Table 1. Description of phenotypic traits and their corresponding Field IDs. Fracture data were obtained from three sources: self-reported questionnaires (*), ICD-10 hospital inpatient records (†), and ICD-9 hospital inpatient records (‡). Heel bone mineral density (heel BMD) was measured via quantitative ultrasound. Abbreviations: ICD, International Classification of Diseases.

Supplementary Table 2. Overview of GWAS and eQTL datasets used in the analysis. All GWAS summary statistics are predominantly based on individuals of European descent unless otherwise specified. Abbreviations: eBMD, estimated bone mineral density; eQTL, expression quantitative trait locus; mvAge, multivariate age.

Supplementary Table 3. Summary of single-cell transcriptomics datasets utilized in this study. Full citations for the original studies are provided.

Supplementary Table 4. Parameters and filtering criteria for two-sample Mendelian Randomization analyses. The table summarized the configuration settings used for each exposure-outcome pair.

Supplementary Table 5. Mean gene expression per cell type for integrated expression atlas. Expression levels were normalized using scVI and scANVI.

Supplementary Table 6. Significant modules identified using scGWAS. Modules with *p* < 0.05, *p_GWAS* < 0.05 and *p_scRNA* < 0.05 were significant. Macrophages were the only cell type harboring modules significantly associated with both traits.

Supplementary Table 7. Significant gene modules identified by scGWAS in macrophage subpopulations. The table lists gene modules significantly associated with aging (mvAge) and bone mineral density (eBMD)

Supplementary Table 8. Gene list of the Gainsboro module. Gainsboro module is enriched for genes whose expression is strongly correlated with macrophages ratios across samples.

Supplementary Table 9. Gene Ontology (GO) enrichment analysis of the Gainsboro gene list obtained by pyWGCNA. Terms are ranked by their Combined Score.

Supplementary Table 10. Gene loci exhibiting a conjunctional false discovery rate (conjFDR) < 0.05 for both mvAge and eBMD, indicating shared genetic associations with these two risk factors.

Supplementary Table 11. Genomic regions showing significant genetic correlation (rho) between mvAge and eBMD. Regions were considered correlated if *p* < 0.05, reflecting shared regulatory pleiotropy.

Supplementary Table 12. Characterization of HNRNPUL1 binding sites on the TGFBR2 transcript. The table details specific RNA-protein interaction events identified via enhanced Cross-Linking and Immunoprecipitation (eCLIP) in HepG2 cells. Data were sourced from the ENCODE project.

